## Supplementary Material for "Comparing Sources of Mobility for Modelling the Epidemic Spread of Zika Virus in Colombia"

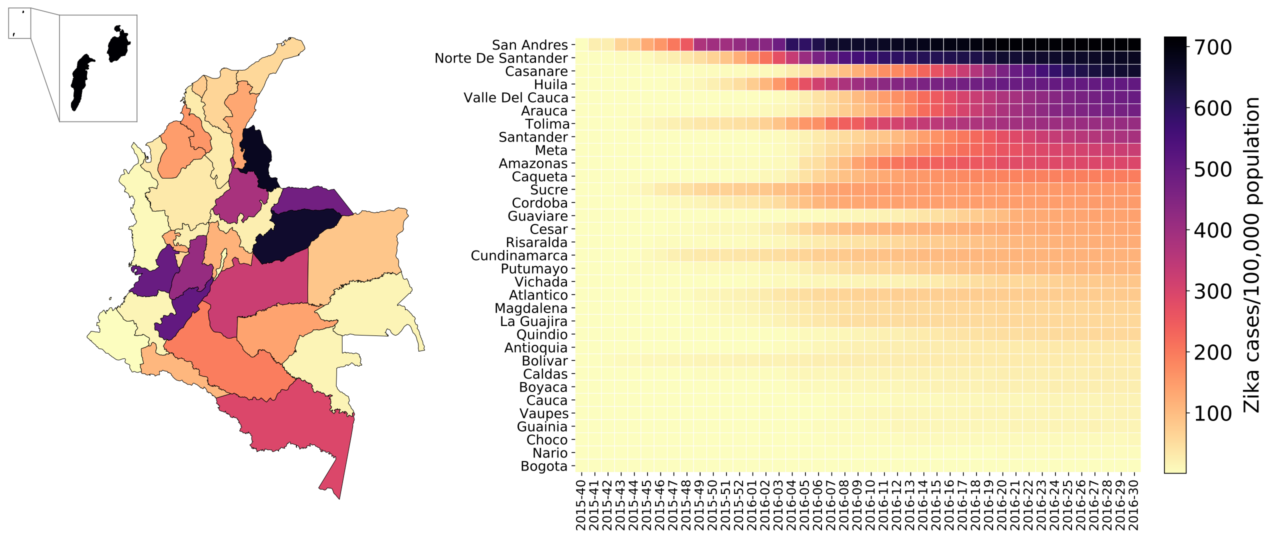


**Fig. S1. Cumulative ZIKV incidence by department in Colombia.** The panels show the cumulative ZIKV incidence (per 100,000 population) by department in the period from October 4, 2015 (epidemiological week 2015-40) to October 2, 2016 (epidemiological week 2016-40) as reported by the epidemiological reports from Colombia's National Institute of Health [1].

### 1 Epidemiological data

The Zika virus outbreak was declared in Colombia in early October 2015, following the identification of the first cluster of laboratory-confirmed cases in nine patients from northern Colombia. With over 100,000 cases reported (of which approximately 8% laboratory confirmed), Colombia had the second highest number of reported cases among the 50 countries with autochthonous transmission during the 2015-2016 outbreak in the Americas.

Here we use the official surveillance data from the Colombia's National Institute of Health (INS) [1] for the entire epidemic period, from the earliest reported cases in epidemiological week 2015-40 to epidemiological week 2016-40. Cases that were retrospectively identified before epidemiological week 2015-32 (beginning on August 9, 2015) are not included because Zika virus disease was not consistently monitored by the nationwide surveillance system before then. The epidemiological reports document the cumulative number of laboratory-confirmed and suspected cases of Zika virus disease by departments and districts. From this, we computed the weekly number of new ZIKV cases by department. The incidence data reported by district was included in the total number for the corresponding department, i.e. the city of Barranquilla in department Atlantico, Buenaventura in Valle Del Cauca, Cartagena in Bolivar, and Santa Marta in Magdalena. Note that the INS did not report the incidence in the Capital District, Bogotá, since the cases originated in other reporting areas. Bogotá was in fact not at risk for autochthonous Zika virus transmission due to the high altitude and low average air temperature which do not favour the presence of mosquitoes [2].

Table S1 reports the population size, the number of Zika virus cases (total and laboratory-confirmed) and the cumulative incidence by department. Figure S1 shows the cumulative ZIKV incidence (per 100,000 population). Note that underreporting due to the clinical similarities of mild symptoms associated with ZIKV, limited diagnostic capabilities, medically unattended cases, and asymptomatic infections (ranging from 50% to 80% [3, 4]), may have contributed significantly to underestimating the actual extent of the epidemic.

**Table S1. Data profiles by department in Colombia.** The table reports the population size of departments and the cumulative number of ZIKV cases (total and laboratory-confirmed) and cumulative ZIKV incidence rates in the period from October 4, 2015 (epidemiological week 2015-40) to October 2, 2016 (epidemiological week 2016-40). Departments are sorted by population size.

| Department | Population (%) | Zika virus cases (confirmed) | Incidence rate (cases/100,000) |
|---|---|---|---|
| Bogota | 7,857,481 (13.74) | 0 (0) | 0 |
| Antioquia | 7,283,785 (12.74) | 2,496 (335) | 34 |
| Valle Del Cauca | 5,050,650 (8.83) | 26,593 (895) | 527 |
| Cundinamarca | 4,487,694 (7.85) | 5,267 (317) | 117 |
| Bolivar | 2,861,055 (5.00) | 1,920 (242) | 67 |
| Atlantico | 2,656,137 (4.65) | 6,765 (359) | 255 |
| Santander | 2,497,070 (4.37) | 10,177 (443) | 408 |
| Cordoba | 2,146,620 (3.75) | 3,220 (253) | 150 |
| Magdalena | 2,063,848 (3.61) | 3,251 (295) | 158 |
| Nario | 1,924,320 (3.37) | 80 (20) | 4 |
| Cauca | 1,707,545 (2.99) | 324 (34) | 19 |
| Tolima | 1,670,891 (2.92) | 7,073 (822) | 423 |
| Norte De Santander | 1,504,770 (2.63) | 10,413 (1,521) | 692 |
| Boyaca | 1,440,444 (2.52) | 364 (88) | 25 |
| Huila | 1,338,588 (2.34) | 6,897 (915) | 515 |
| La Guajira | 1,230,463 (2.15) | 721 (95) | 59 |
| Meta | 1,229,986 (2.15) | 4,273 (580) | 347 |
| Cesar | 1,174,300 (2.05) | 1,611 (245) | 137 |
| Caldas | 1,071,772 (1.87) | 303 (74) | 28 |
| Risaralda | 1,041,744 (1.82) | 1,297 (130) | 125 |
| Sucre | 1,014,279 (1.77) | 1,627 (107) | 160 |
| Quindio | 662,152 (1.16) | 394 (24) | 60 |
| Caqueta | 588,279 (1.03) | 1,127 (234) | 192 |
| Choco | 585,044 (1.02) | 57 (5) | 10 |
| Casanare | 584,037 (1.02) | 3,912 (280) | 670 |
| Putumayo | 468,906 (0.82) | 527 (110) | 112 |
| Arauca | 381,776 (0.67) | 1,848 (191) | 484 |
| San Andres | 157,011 (0.27) | 1,142 (66) | 727 |
| Guaviare | 146,890 (0.26) | 211 (15) | 144 |
| Amazonas | 112,449 (0.20) | 330 (28) | 293 |
| Vichada | 87,116 (0.15) | 76 (5) | 87 |
| Guainia | 78,773 (0.14) | 14 (3) | 18 |
| Vaupes | 76,603 (0.13) | 14 (0) | 18 |
| Colombia | 57,182,478 (100) | 104,324 (8,731) | 183 |

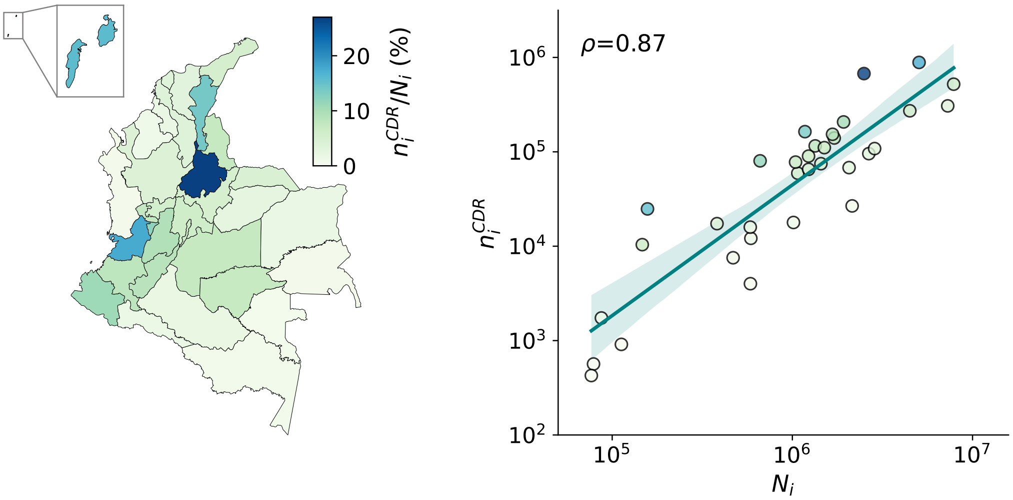


**Fig. S2. Population sampling ratio in the CDR-informed network.** The map shows the population sampling ratio $n_i^{CDR}/N_i$ by department (left) and the relationship between the CDR-derived population sample $n_i^{CDR}$ and the resident population $N_i$ (right), that show good agreement with Spearman correlation $\rho$=0.87. The colour code refers to the population sampling ratio.

### 2 CDR-informed network

CDR-derived movements are identified when the same phone number is handled by two different cell towers in two consecutive calls. Here we use aggregated CDR-derived movements obtained from more than two billion encripted and anonymized metadata calls made over a six-month period, from December 2, 2013 to May 19, 2014 (see Coscia et al. [5] for more details). The data consists of weekly OD matrices of number of trips $T_{ij}^w$ from municipality $i$ to municipality $j$ occurred in week $w$ and weekly number of active phone numbers $n_i^w$ in municipality $i$ in week $w$, where $w$ goes from calendar week 2013-49 to calendar week 2014-21. Note that this data therefore does not refer to daily commuting patterns based on users' most frequently visited locations. However, given the long observation period and large operator coverage, we assume that potential variability due to long-distance travels, weekly and/or seasonal fluctuations, major vacation periods, etc., are smoothed when considering average values.

To generate the CDR-informed mobility network, $w_{ij}^{CDR}$, we follow the next steps. First, we spatially aggregate values at the level of departments. Note that in this operation most of the trips $T_{ij}^w$ are lost during this operation as they inevitably end up in the same department. Figure S3 shows the fluctuations in the number of active phones $n_i^w$ and trips $T_{ij}^w$ by department (sorted by mean active phones). The data covers all 33 Colombian departments, apart from the department of Caqueta during weeks 2013-49, 2013-50, 2013-51. The number of directed links (i,j) ranges from a minimum of 676 links to a maximum of 845 links. On average, there are over 900,00 trips occurred in a week, ranging from 746,350 to 1,102,646 trips, and over 4 million active phone numbers in a week, corresponding to approximately 9% of the total population in Colombia.

Secondly, we normalize flows in order to correct for potential biases due to under- and/or over-sampling and to match the same population size. This is because CDR-derived mobility is inevitably biased by population sampling and coverage. Cell towers are typically distributed according to population density, but mobile phone activity is heterogeneous across space. Here

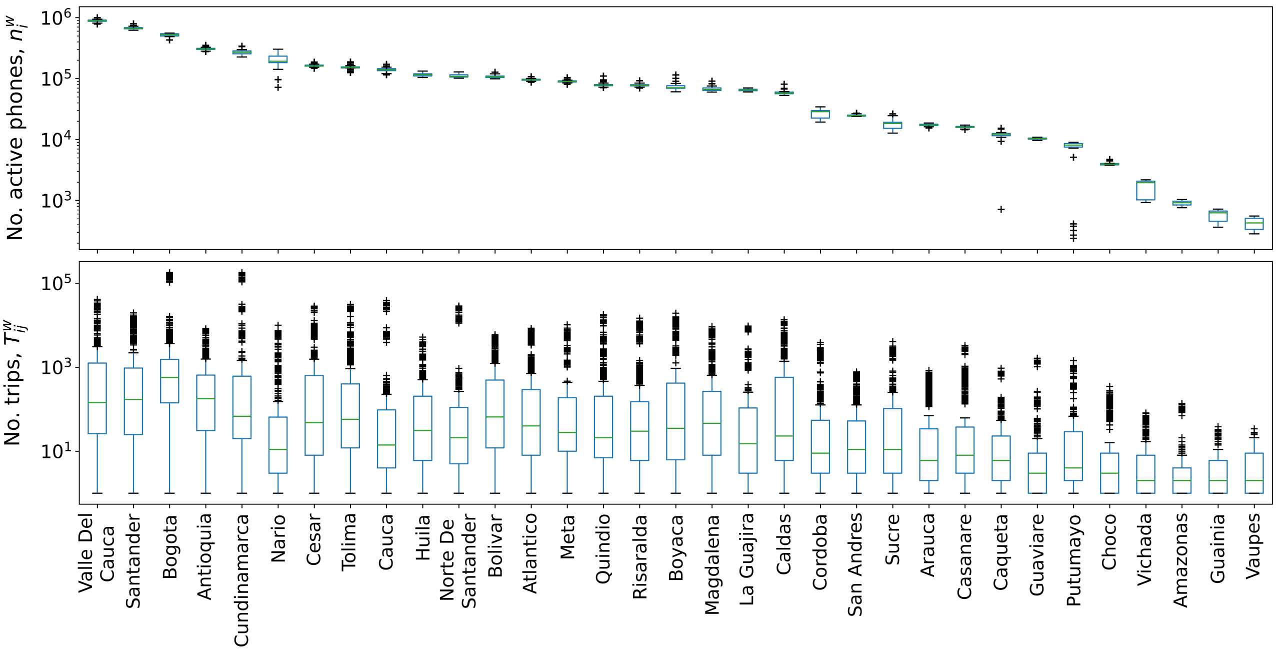


**Fig. S3. Variability in the CDR-derived mobility dataset.** The boxplots show the fluctuations in the number of active phones $n_i^w$ (top) and the number of trips $T_{ij}^w$ (bottom). Departments on the x-axis are sorted according to the mean active phones.

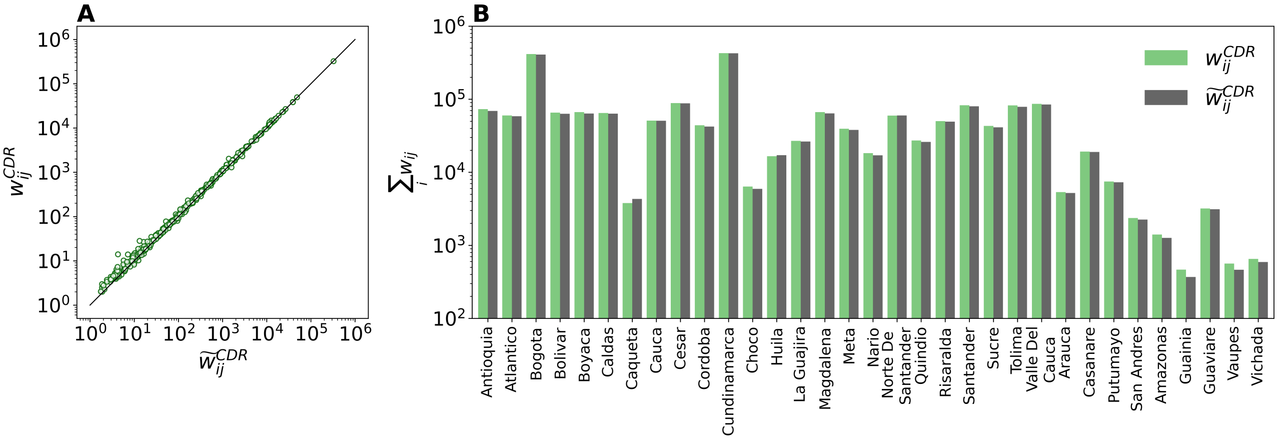


**Fig. S4. Sensitivity analysis on the CDR-informed network.** Comparison between the flows $w_{ij}^{CDR}$ and the flows $\widetilde{w_{ij}}^{CDR}$ calculated as the median normalized flows over the weeks. (**A**) Relationship between the flows $w_{ij}^{CDR}$ and $\widetilde{w_{ij}}^{CDR}$. (**B**) Comparison between the outflows $\sum_i w_{ij}^{CDR}$ and $\sum_i \widetilde{w_{ij}}^{CDR}$ by department.

we compute weights based on the population sampling ratio $n_i^w/N_i$ at origin $i$ with resident population $N_i$ and compute the normalized flows as $w_{ij}^w = T_{ij}^w N_i/n_i^w$ (see Figure S2).

Lastly, we average flows over the weeks $w$ (missing values in a given week are treated as null values, i.e. no trips were observed during that week) and those connections having $w_{ij} < 1$. As a sensitivity analysis, we create an alternative network by computing the median flows over the weeks $w$ (indicated as $\widetilde{w_{ij}}^{CDR}$). This allows us to flatten those unusually high flows or sporadic connections due to major events, such as Christmas. However, we found no substantial difference between the two networks (Spearman's $\rho$=0.99), as shown in Figure S4.

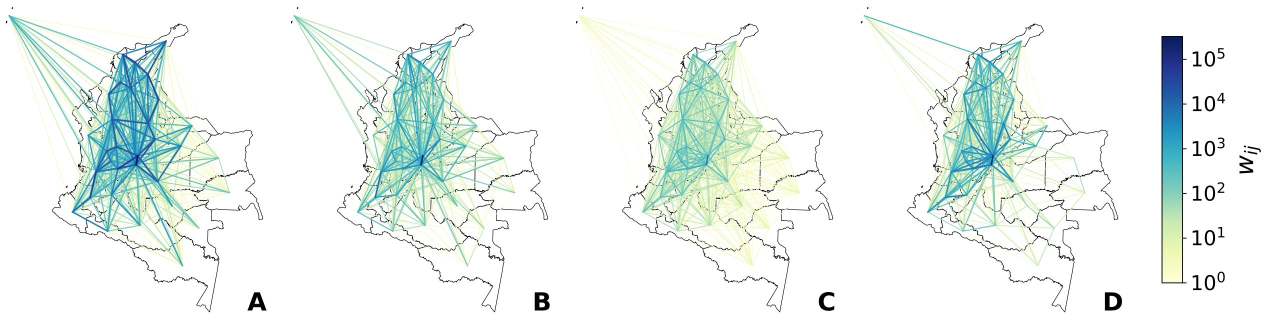


**Fig. S5. Mobility networks.** The maps shows the flows $w_{ij}$ of individuals among departments of Colombia in the CDR-informed network (**A**), the census network (**B**), the gravity network (**C**), and the radiation network (**D**). The solid lines correspond to the links, while the colour code represents the weights.

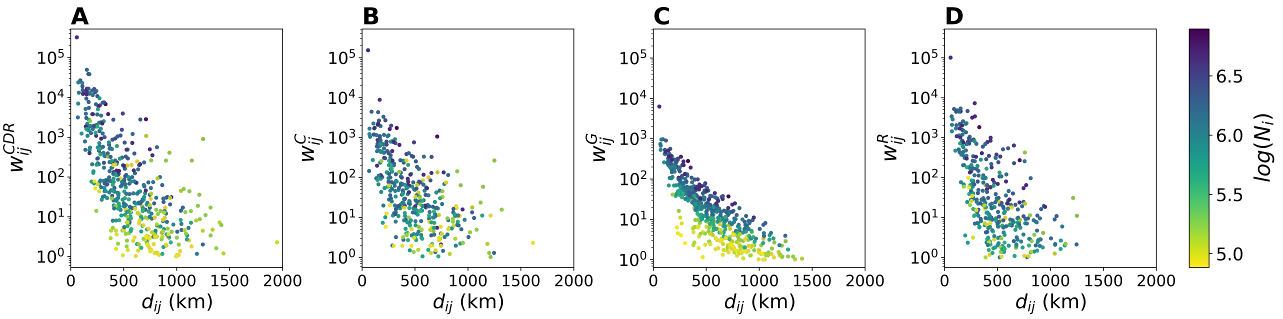


**Fig. S6. Mobility flows by distance and population size.** The plots show the relationship between the flows $w_{ij}$ (y-axis) and the distances $d_{ij}$ calculated between centroids (x-axis) for the CDR-informed network (**A**), the census network (**B**), the gravity network (**C**), and the radiation network (**D**). The colour code corresponds to the population size $N_i$ at origin $i$ (log scale).

### 3 Gravity model

As described in the main text, we use the gravity model [6] to synthetically infer the flows $w_{ij}$ of individuals between different locations according to Eq. (1). The gravity model requires previously existing traffic data in order to fit the free parameters, i.e. the proportionality constant $C$, the exponents $\alpha$ and $\gamma$ of the population sizes at origin and destination, and the constant $\beta$ that tunes the distance dependence. In particular, the distance-dependent function typically follows a power or exponential law depending on the geographic resolution of the study [refs]. Here we tested both functional forms, i.e. $f(d_{ij}) = d_{ij}^{\beta}$ and $f(d_{ij}) = e^{\beta d_{ij}}$, respectively. Distances $d_{ij}$ are here calculated between the centroids of locations $i$ and $j$ using the Vincenty's formula. Following the approach of Balcan et al. [6], we apply a multivariate linear regression analysis in the logarithmic scale to estimate the best functional form for $f(d_{ij})$ and the values of the free parameters that best fit the fluxes $w_{ij}$ of the census data.

We found the best performance using a power distance-decay function. The regression coefficients are $R^2 = 0.55$ and $R^2 = 0.46$ respectively for the power and the exponential distance-decay function. Table S2 reports the estimated values for $\alpha$, $\gamma$, $\beta$ and $C$. All p-values are statistically significant, except for the intercept ($p = 0.7$), meaning that we cannot reject the null hypothesis that the coefficient is equal to 0, thus yielding $C = 1$.

**Table S2. Parameters of the gravity model.** Estimated values of the free parameters in the gravity model as obtained by applying a multivariate linear regression to the empirical census data.

| Parameter | Estimate | Standard Error | p-value |
|---|---|---|---|
| $\alpha$ | 0.60 | 0.06 | $< 10^{-26}$ |
| $\gamma$ | 0.46 | 0.05 | $< 10^{-20}$ |
| $\beta$ | 1.93 | 0.10 | $< 10^{-75}$ |
| $C$ | 1.43 | 3.38 | 0.3 |

Given the values of the parameters, we apply the gravity law of Eq. (1) to generate the mobility fluxes $w_{ij}^{G}$. We obtain the final synthetic mobility network by symmetrizing the fluxes and removing those connections having $w_{ij} < 1$.

### 4 Modelling the epidemic spread of ZIKV

The infection dynamics occurs in homogeneous mixing approximation within each subpopulation according to a compartmental classification of the individuals based on the various stages of the disease. Humans can occupy one of four compartments: susceptible individuals $S_H$ who lack immunity against the infection, exposed individuals $E_H$ who have acquired the infection but are not yet infectious, infected individuals $I_H$ who can transmit the infection (and may or may not display symptoms), and removed individuals $R_H$ who no longer have the infection and are immune to further ZIKV infection. Since the average human lifespan is much longer than the outbreak duration, we omit human births and deaths and we consider human population size constant, i.e. $S_H + E_H + I_H + R_H = N_H$. Susceptible humans transition to the exposed compartment occurs under the vector-to-human force of infection that follows the usual mass-action law and is given by the expression $\lambda_{VH} = \beta\tau_{VH}\frac{I_V}{N_H}$, where $\beta$ accounts for the daily mosquito biting rate and the specific transmissibility of ZIKV, and $\tau_{VH}$ corresponds to the mosquito-to-human probability of transmission. The mosquito-to-human probability of transmission $\tau_{VH}$ is temperature-dependent and follows the expression [7]:

$$\tau_{VH} = 0.001044T(T - 12.286)\sqrt{32.461 - T} \quad (1)$$

Exposed individuals become infectious at a rate $\epsilon_H$, which is inversely proportional to the mean intrinsic latent period of the infection, $\epsilon_H^{-1}$. Infectious individuals then recover from the disease at a rate $\mu_H$, which is inversely proportional to the mean infectious period, $\mu_H^{-1}$. On average, individuals remain in the exposed and infectious compartment for the duration of the mean intrinsic latent period $\epsilon_H^{-1}$=7 days and the mean infectious period $\mu_H^{-1}$=5 days, respectively.

The mosquito vector population is described by the number of susceptible $S_V$, exposed $E_V$, and infectious mosquitoes $I_V$. The number of mosquitoes per person is expressed by $k$, so that $k = \frac{N_V}{N_H}$. It is temperature-dependent and follows a modulation function given by [8]:

$$k \propto exp(-(\tilde{T}(t) - 25)^2/50) \quad (2)$$

where $\tilde{T} = \sum_{i=0}^{78} T(t-1)/79$ is the average temperature of the previous 79 days, where $t$ is the time in days and $T(t)$ is the average temperature on day $t$.

The human-to-vector force of infection $\lambda_{HV}$ governing the transition rate from susceptible to exposed individuals is given by $\lambda_{HV} = \beta\tau_{HV}\frac{I_V}{N_H}$, where $\tau_{HV}$ corresponds to the human-to-mosquito probability of transmission. Exposed mosquitoes transition to the infectious class occurs at a rate $\epsilon_V$, which is inversely proportional to the mean extrinsic latent period in the mosquito population, $\epsilon_V^{-1}$. Mosquitoes in each compartment die at a rate $\mu_V$, that is inversely proportional to the mosquito lifespan, $\mu_V^{-1}$, and are re-introduced in the susceptible compartment at the same rate to allow the replenishment of mosquitoes after death. On average, mosquitoes remain in the exposed compartment for the duration of the mean extrinsic latent period $\epsilon_V^{-1}$=8 days. The average lifetime of mosquitoes $\mu_V^{-1}$ is instead temperature-dependent and follows the expression [9]:

$$\mu_V(T) = 0.3967 - 0.03912T + 2.442 \times 10^{-3}T^2 - 7.479 \times 10^{-5}T^3 + 9.298 \times 10^{-7}T^4 \quad (3)$$

The basic reproduction number $R_0$ can be expressed following the standard approach of Ref. [10] and thus varies spatio-temporally according to the temperature and mosquito abundance:

$$R_0 = \frac{\epsilon_V}{\mu_H\mu_V(\epsilon_V + \mu_V)}k\beta^2\tau_{VH}\tau_{HV} \quad (4)$$

The model is fully stochastic and individuals are assumed to be homogeneously mixed within each subpopulation. The coupled population equations describing the epidemic time evolution read as follows:

$$\begin{aligned}
S^H_{t+1} &= S^H_t - \Delta_{S^H \longrightarrow E^H} \\
E^H_{t+1} &= E^H_t + \Delta_{S^H \longrightarrow E^H} - \Delta_{E^H \longrightarrow I^H} \\
I^H_{t+1} &= I^H_t + \Delta_{E^H \longrightarrow I^H} - \Delta_{I^H \longrightarrow R^H} \\
R^H_{t+1} &= R^H_t + \Delta_{I^H \longrightarrow R^H}, \\
S^V_{t+1} &= S^V_t - \Delta_{S^V \longrightarrow E^V} + \Delta_{E^V \longrightarrow S^V} + \Delta_{I^V \longrightarrow S^V} \\
E^V_{t+1} &= E^V_t + \Delta_{S^V \longrightarrow E^V} - \Delta_{E^H \longrightarrow S^V} - \Delta_{E^V \longrightarrow I^V} \\
I^V_{t+1} &= I^V_t + \Delta_{E^V \longrightarrow I^V} - \Delta_{I^V \longrightarrow S^V}.
\end{aligned} \quad (5)$$

Each term $\Delta_{X \longrightarrow Y}$ represents the number of human or vector individuals transitioning from state $X$ to state $Y$. Transitions are simulated through chain binomial processes $\Delta_{X \longrightarrow Y} = Binomial(X, p_{X \longrightarrow Y})$ considering a number of trials equal to the number of individuals $X$ in the compartments and a transition probability $p_{X \longrightarrow Y}$ determined by the force of infection and the average lifetime of individuals in each compartment. Each transition is a memoryless discrete stochastic transition process.

#### 4.1 GLEAM for ZIKV analysis

In this work, we adopt the calibration of the ZIKV transmission model from Zhang et al. [8] on the use of the Global Epidemic And Mobility model (GLEAM) to simulate the 2015-2016 ZIKV epidemic in the Americas. GLEAM is a stochastic generative model that leverages real world data to perform in-silico simulations of the spatial spread of infectious diseases at the global level. Its validity has been shown in several historical and ongoing outbreaks, such as the 2009 H1N1 influenza pandemic [11, 12], Ebola in West Africa [13], Zika virus disease in the Americas [8], and the COVID-19 pandemic [14].

In more detail, GLEAM is based on a metapopulation approach whose subpopulations are defined through a Voronoi tessellation of the Earth surface in cells of approximately 25×25km square (along Earth's equator), which are grouped around the major transportation hubs. This results into over 3,200 subpopulations connected by a network of travel fluxes that includes both short-range mobility (commuting) and long-range mobility based on the origin-destination matrices of global air travel [ref]. Within each subpopulation, a compartmental model is used to simulate the disease of interest, whose transitions are stochastically defined by chain binomial and multinomial processes. Population data is obtained from the database of the Gridded Population of the World project from the Socioeconomic Data and Application Center at Columbia University (SEDAC), consisting of population estimates in 2015 per grid-cell 1kmx1km (`sedac.ciesin.columbia.edu`).

In the analysis of ZIKV, GLEAM integrates additional data layers to account for the spatial heterogeneity driven by the presence of the vector and the characteristics of the exposed populations due to socio-economic factors. This is because sustained local transmission of ZIKV is possible only in those areas where the local environment and climate favour the proliferation of mosquitoes [2], but at the same, even when the environmental conditions are suitable for the transmission of ZIKV, the socio-economic factors modulate the exposure of the population to the vector itself. To this end, GLEAM integrates:

- The global air temperature dataset [15] containing monthly average temperatures at a spatial resolution of $0.5° \times 0.5°$. To match the spatial resolution of GLEAM's gridded population density map, the temperature for each population cell is extracted from the nearest available point in the temperature dataset. Daily average temperatures are linearly interpolated from each population's monthly averages.

- The global distribution of *Aedes* aegypti and *Aedes* albopictus at a spatial resolution of 5km × 5km [16]

- The geophysically scaled economic dataset (G-Econ) developed by Nordhaus et.al. [17, 18] containing the per capita Gross Domestic Product (computed at Purchasing Power Parity exchange rates) at a spatial resolution of $1° \times 1°$. To estimate the per capita Gross Cell Product (GCP) at Purchasing Power Parity (PPP) rates, the amount is distributed across GLEAM cells proportionally to each cell's population size. The data has also been rescaled to reflect 2015 GDP per capita (PPP) estimates.

These datasets are combined to model the key drivers of ZIKV transmission. Temperature is used to compute many important disease parameters and to define the daily mosquito abundance (number of mosquitoes per human) in each cell. Data on mosquito abundance and temperature are used to identify cells where ZIKV outbreaks are not possible because of environmental factors. The human populations in these cells are thus considered unexposed to ZIKV. Finally, historical data and G-Econ are used to provide a socio-economic rescaling factor, reflecting how exposure to the vector is impacted by socioeconomic variables such as the availability of air-conditioning.

The calibration of the ZIKV transmission model is performed by a Markov Chain Monte Carlo (MCMC) analysis of surveillance data reported from the 2013 ZIKV epidemic in French Polynesia [19]. Setting the extrinsic and intrinsic latent periods and the human infectious period to reference values and using average daily temperatures of French Polynesia, we estimate a basic reproduction number at the temperature T = 25 C° from French Polynesia as RFP 0 = 2.75 (95% CI [2.53-2.98]), which is consistent with other ZIKV outbreak analyses [19, 20]. The calibration in French Polynesia provides the basic transmissibility of ZIKV.

Once the data layers and parameters have been defined, the model runs using discrete time steps of one full day to simulate the transmission dynamic model, incorporating human mobility between subpopulations. The model is fully stochastic and from any nominally identical initialization (initial conditions and disease model), generates an ensemble of possible epidemics, as described by newly generated infections, time of arrival of the infection in each subpopulation, and the number of traveling carriers. The Latin square sampling of the initial introduction of ZIKV in the Americas, and the ensuing statistical analysis is performed on 150,000 stochastic epidemic realizations. The probability $p(x)$ and $p(x|\theta)$ computed from those realizations are defined as the probability of the evidence (the epidemic peak in Colombia, as from surveillance data) and the likelihood of the evidence given the parameters $\theta$ specifying the date and location of introduction of ZIKV in Brazil. The posterior probabilities of interest are computed accordingly from those distributions. In total, 1736 out of the 150,000 simulations match the epidemic peak in Colombia and the posterior estimate on the time of introduction into Brazil matches phylogenetic evidence [21, 22]. The ensemble of the 1736 simulations are therefore considered as the output of the global model to characterize the spatiotemporal dynamics of ZIKV transmission in the Americas and the global dissemination of the travel related ZIKV infections through human mobility.

Further details and the full description of GLEAM in the analysis of the epidemic spread of Zika virus in the Americas can be found in Zhang et al. [8].

#### 4.2 ZIKV importation cases to Colombia

In this work, we use the simulation output of GLEAM on the introduction of ZIKV infections into Colombia [8]. This corresponds to a total of 1,189 simulations of ZIKV infections imported into Colombia for which we know the time of arrival, the stage of ZIKV infection (exposed or infectious), and the airport of origin and arrival. At each iteration in the model, we randomly sample one of the 1,189 simulations of imported ZIKV infections and we use it to feed the compartments of exposed $E_H$ or infectious $I_H$ individuals in the corresponding subpopulation of arrival.

Figure S7A shows the time-series boxplot of the daily ZIKV infections imported into Colombia in the period between January 1, 2015 and December 31, 2016. In agreement to the genetic findings, first ZIKV cases arrived in Colombia between January and April 2015 [23]. The total number of ZIKV cases imported into Colombia has a median value of 8,671 cases (IQR: 8,315-9,064), while the number of ZIKV cases imported daily has a median value of 10 cases (IQR: 3-21).

Figures S7B and S7C show the imported ZIKV infections by country of origin and department of arrival in Colombia. The countries that contribute the most to the importation of Zika virus cases into Colombia are Brazil and Panama with an average of 1,747 ZIKV imported cases (IQR: 1,660-1,835) and 1,724 (IQR: 1,502-1,946), respectively, corresponding to approximately 40% of the total imported cases. Other major contributing countries are Ecuador (about 12%),Venezuela (about 11%),and Mexico (about 9%). Conversely, the most affected department is the Capital District Bogotá with an average of 3,070 imported ZIKV cases (IQR: 2,940-3,211), corresponding to approximately 35% of the total imported cases. On average, the ZIKV cases imported into Bogotá daily is 4 (IQR: 2-8). Other departments most affected are Cundinamarca with 1,953 cases (IQR: 1,868-2,042) (about 23%), Antioquia with 1,112 (IQR: 1051-1,177) (about 13%), Valle del Cauca 825 (IQR: 779-872) (about 10%), and Bolivar 357 (IQR: 336-381) (about 4%).

The same rescaling factor due to environmental and socio-economic conditions applies to the

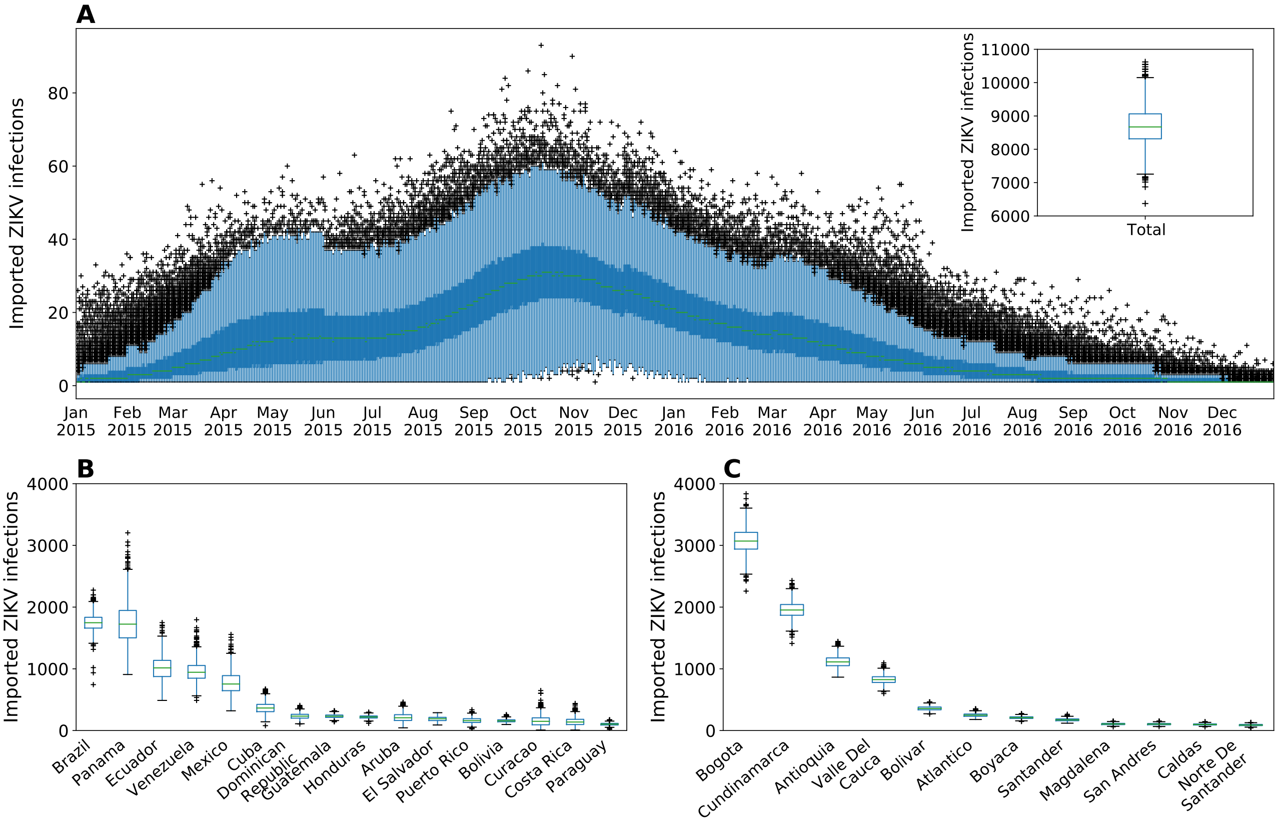


**Fig. S7. Imported ZIKV infections into Colombia.** (**A**) Time-series boxplot of total ZIKV cases imported daily into Colombia. (**B**) Imported ZIKV cases by country of origin. (**C**) Imported ZIKV cases by department of arrival in Colombia.

imported ZIKV infections such that the likehood of seeding an epidemic locally varies depending on whether the subpopulation of destination is at risk or not of ZIKV transmission. Figures S8 shows the average number of ZIKV infections imported daily into the whole population and the exposed population in the departments of Bogotá (A), Cundinamarca, (B) Antioquia (C) and Valle del Cauca (D). Although the importation of ZIKV infections interests mostly Bogotá, environmental and socio-economic conditions do not favour local epidemic. On the other hand, these individuals will eventually travel and spread the virus in other regions more favourable to ZIKV transmission.

### 5 Statistical analysis

We compare the structural properties of the various mobility network with the census network. To this end, we restrict the analysis to their topological intersection and perform various statistical and similarity measures, including the Kendall's $\tau$ and Spearman's $\rho$ correlation coefficients (computed both on flows $w_{ij}$ and outflows $\sum_i w_{ij}$), the Jaccard index, the cosine similarity, and the common part of commuters (CPC). We considered p-values below 0.01 to be statistically significant.

Let's consider a network $A$ with elements $A_{ij}$ and weights $w_{ij,A}$, and a network $B$ with elements $B_{ij}$ and weights $w_{ij,B}$. The Jaccard index measures the proportion of shared links between $A$ and $B$ relative to the total number of links in $A$ and $B$. The Jaccard index is defined as follows:

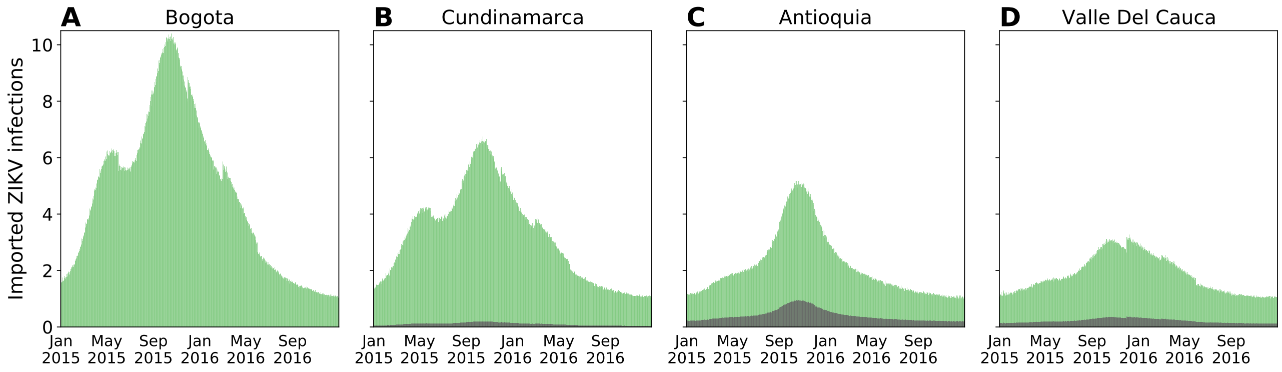


**Fig. S8. Imported ZIKV infections by department of arrival.** Average daily number of ZIKV infections imported into the whole population (green) and into the exposed population (grey) for the departments of Bogotá (A), Cundinamarca (B), Antioquia (C), and Valle del Cauca (D).

$$J(A_{ij}, B_{ij}) = \frac{|A_{ij} \cap B_{ij}|}{|A_{ij} \cup B_{ij}|}, \quad J(A_{ij}, B_{ij}) \in [0, 1] \tag{6}$$

Values of $J$ close to 1 indicate that the two networks share a large fraction of links, while values close to 0 indicate that the two networks have very different structures.

The cosine similarity considers both links and weights shared by two network and is defined as follows:

$$\sigma(w_{ij,A}, w_{ij,B}) = \frac{\sum_j w_{ij,A} w_{ij,B}}{\sqrt{\sum_j w_{ij,A}^2 \sum_j w_{ij,B}^2}}, \quad \sigma(w_{ij,A}, w_{ij,B}) \in [0, 1] \tag{7}$$

It takes values from 0, i.e. the sets of links are completely different in the two configurations, to 1, i.e. the two networks share the same set of links with same weights.

The common part of commuters (CPC) is a measure of similarity (based on the Sørensen-Dice similarity coefficient) that compares the weights according to the following expression:

$$CPC(w_{ij,A}, w_{ij,B}) = \frac{2 \sum_{i,j} min(w_{ij,A}, w_{ij,B})}{\sum_{i,j} w_{ij,A} + \sum_{i,j} w_{ij,B}} \tag{8}$$

Values of $CPC(w_{ij,A}, w_{ij,B})$ varies from 0, when no agreement is found, to 1, when the two networks are identical.

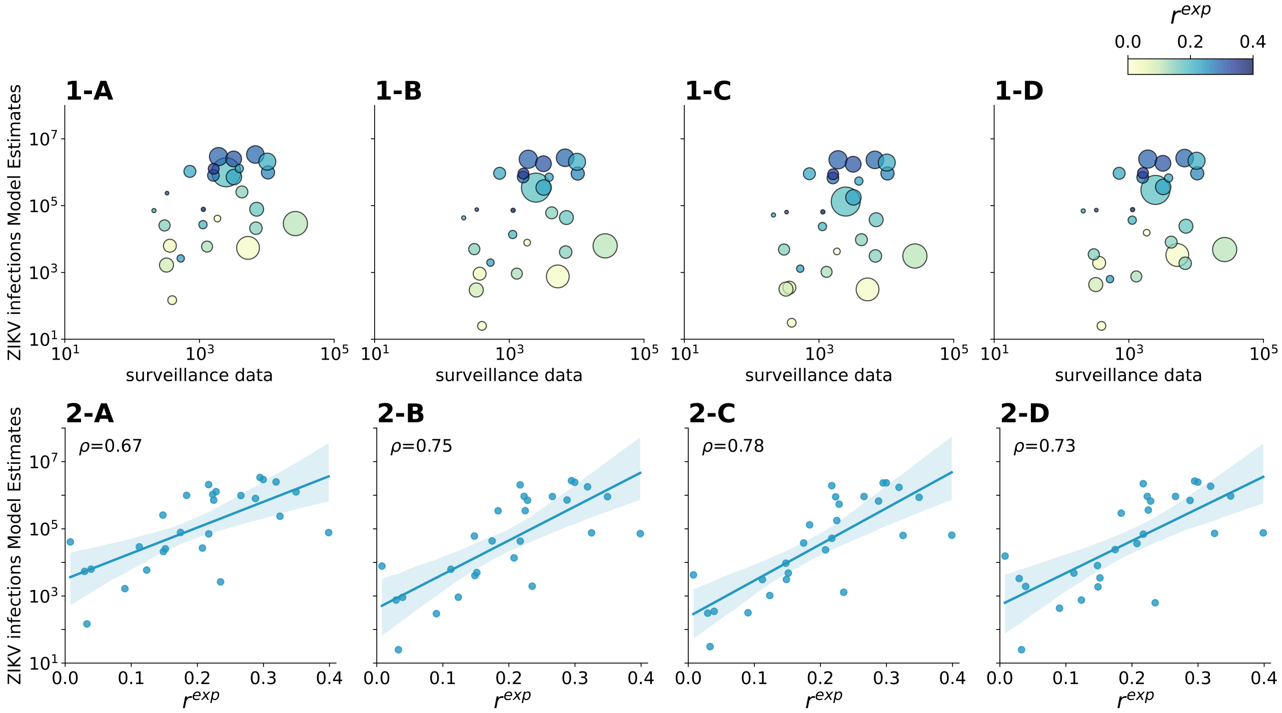


**Fig. S9. The impact of population exposure on the epidemic outcome.** The top panel shows the comparison between the median values of the model estimates (y-axis) and the official surveillance data (x-axis) by department. Points size correspond to population size. Colour code correspond to the rescaling factor $r^{exp}$, that is the fraction of the population exposed to the virus due to environmental and socio-economic conditions. The bottom panel shows the correlation between the model estimates (y-axis) and the rescaling factor $r^{exp}$ (x-axis). Spearman's $\rho$ correlation coefficients are reported for the linear association. Columns correspond to the mobility networks, i.e. the CDR-informed network (A), the census network (B), the gravity network (C), and the radiation network (D).
